# The GenoPred Pipeline: A Comprehensive and Scalable Pipeline for Polygenic Scoring

**DOI:** 10.1101/2024.06.12.24308843

**Authors:** Oliver Pain, Ammar Al-Chalabi, Cathryn M. Lewis

## Abstract

**Motivation:** Polygenic scoring is a commonly used approach for estimating an individual’s likelihood of a given outcome. Polygenic scores are typically calculated using genetic effects derived from genome-wide association study (GWAS) summary statistics and individual-level genotype data for the target sample. Using a reference-standardised framework ensures the polygenic score can be reliably interpreted. Going from genotype to interpretable polygenic scores involves many steps and there are many methods available, limiting the accessibility of polygenic scores for research and clinical application. Additional challenges exist for studies in ancestrally diverse populations. We have implemented the leading polygenic scoring methodologies within an easy-to-use pipeline called GenoPred.

**Results:** Here we present the GenoPred pipeline, an easy-to-use, high-performance, reference-standardised and reproducible workflow for polygenic scoring. The pipeline requires just a few readily available inputs to get started, with configuration options available to cater for a range of use-cases. GenoPred implements a comprehensive set of analyses, including genotype and GWAS quality control, target sample ancestry inference, polygenic score file generation using a range of leading methods, and target sample scoring. GenoPred standardises the polygenic scoring process using reference genetic data, providing interpretable polygenic scores, and improving the transferability of results to external datasets. The pipeline is applicable to GWAS and target data from any population within the reference, facilitating studies of diverse ancestry. GenoPred is a Snakemake pipeline with associated Conda software environments, ensuring reproducibility. We apply the pipeline to UK Biobank data demonstrating the pipeline’s simplicity, efficiency, and performance. GenoPred is open-source software, that will continue to develop as polygenic scoring methodology develops.

**Conclusions:** The GenoPred pipeline provides a novel resource for polygenic scoring, integrating a range of complex processes within an easy-to-use framework. GenoPred widens access of the leading polygenic scoring methodology and their application to studies of diverse ancestry.

## Introduction

The advent of genome-wide association studies (GWAS) has revolutionised our understanding of the genetic architecture of complex traits and diseases. GWAS identify associations between genetic variants and traits across the genome, providing valuable insights into the biological pathways influencing disease risk and trait variability (Adams et al., 2024; Van Rheenen et al., 2021; Yengo et al., 2022). A key application of GWAS findings is the development of polygenic scores, which aggregate the effects of numerous genetic variants across the genome to estimate an individual’s genetic predisposition to a given trait or disease (Choi et al., 2020; Dudbridge, 2013). Polygenic scoring has shown promise in predicting disease risk (Khera et al., 2018), informing personalized medicine (Fahed et al., 2022), and contributing to the understanding of genetic overlap between phenotypes (Pain et al., 2018; Pain, Hodgson, et al., 2022; Power et al., 2015).

Despite their potential, the application of polygenic scores in both research and clinical settings faces significant challenges. The process of calculating polygenic scores from genotype data and GWAS summary statistics involves multiple complex steps, including quality control, ancestry inference, generation polygenic scoring files, and target sample scoring (Choi et al., 2020; Pain et al., 2021). Each of these steps is crucial for the calculation of reliable and interpretable polygenic scores. However, the diversity of methodologies makes it difficult for researchers and clinicians to adopt polygenic scoring widely. This complexity limits the accessibility of polygenic scoring and its potential benefits for understanding and predicting complex traits and diseases.

Recognizing these challenges, we introduce the GenoPred pipeline, a comprehensive, easy-to-use pipeline designed to streamline the polygenic scoring process. The pipeline automates and extends a workflow that we used to compare polygenic scoring methods (Pain et al., 2021), integrating leading polygenic scoring methodologies within a reference-standardised framework, facilitating the generation of interpretable and transferable polygenic scores. By simplifying and standardising the polygenic scoring process, the GenoPred pipeline aims to enhance the accessibility and application of polygenic scores across diverse research and clinical contexts.

This paper presents the GenoPred pipeline, detailing its features, implementation, and application to real-world data. We demonstrate GenoPred’s performance and utility by applying it to UK Biobank, showcasing its ability to produce reliable polygenic scores, in a computationally efficient manner. As an open-source tool, GenoPred can evolve alongside advancements in polygenic scoring methodologies, ensuring its continued relevance and utility in the field.

In the following sections, we describe the design and functionality of the GenoPred pipeline, demonstrate its performance through application to UK Biobank data, and discuss the implications of our work for the future of polygenic scoring in both research and clinical applications.

### Overview of GenoPred Pipeline

The GenoPred pipeline provides a sophisticated workflow designed to facilitate polygenic scoring, leveraging the robustness of Snakemake - a workflow management system - and Conda for software environment management (Mölder et al., 2021). This pipeline is engineered to utilize multiple cores for parallel processing and is adaptable for deployment across High-Performance Computing (HPC) or cloud computing systems, ensuring scalability and flexibility in research applications. We also provide docker and singularity software containers to run the GenoPred pipeline, facilitating the pipeline’s use, particularly within offline environments, where no internet is available when accessing secure data.

#### Input Data Processing

GenoPred accepts a range of inputs formats, minimising file preparation in advance of running the pipeline. Individual-level genotype data for the target samples, in which polygenic scores are to be calculated, can be provided in PLINK1, PLINK2, BGEN, VCF or 23andMe format. GWAS summary statistics can have a range of headers, with GenoPred automatically interpreting them. GenoPred automatically detects the genome build of input data, allowing for builds GRCh36 (hg18), GRCh37 (hg19) or GRCh38. If chromosome and base pair information is present, RSIDs are automatically inserted by GenoPred. All inputs are harmonised with the reference genetic data, removing variants that cannot be identified in the reference, updating RSIDs, and resolving strand flips. GenoPred also has the ability to use externally provided score files containing genetic effects to be used for polygenic scoring. The user can specify Polygenic Score Catalog IDs, which GenoPred will automatically download, or the user can provide locally-stored score files following the Polygenic Score Catalog header format (https://www.pgscatalog.org/downloads/#scoring_header)(Lambert et al., 2021).

#### Ancestry Inference

It is important to take into consideration an individual’s ancestry when analysing and interpreting their polygenic scores. GenoPred includes an ancestry inference step, matching target individuals to populations present in the reference dataset. By default, the reference data set is a combination of samples from 1000 Genomes phase 3 (1KG)(1000 Genomes Project Consortium, 2015) and the Human Genome Diversity Project (HGDP)(Bergström et al., 2020), capturing a range of global populations, though the user can specify alternative references to be used. The ancestry inference results enable the scaling of polygenic scores according to an ancestry-matched reference, standardising scores to units of standard deviation from the mean of the matched reference population.

#### Leading Polygenic Scoring Methodology

GenoPred integrates seven leading polygenic scoring methods that adjust GWAS effects sizes for polygenic scoring. These include *p*-value thresholding and clumping (pt+clump)(Chang et al., 2015), lassosum (Mak et al., 2017), DBSLMM (Yang & Zhou, 2020), LDpred2 (Privé et al., 2020), SBayesR (Lloyd-Jones et al., 2019), MegaPRS (Zhang et al., 2021), and PRS-CS (Ge et al., 2019). The default parameters used for each method are shown in the technical documentation (Supplementary Data File 1). Certain parameters for each method can be altered by the user, such as selecting only the ‘auto’ model within LDpred2. This diversity in methodologies ensures that users can select the approach best suited to their study’s needs. The adjusted genetic effects, referred to as score files, are then used to calculate polygenic scores in target samples using PLINK2 (Chang et al., 2015).

#### Principal Component Estimation

GenoPred enables calculation of both within-sample or reference-projected principal components (PCs). Within-sample PCs are estimated using the target sample data, which are commonly included as covariates when performing association analyses (Price et al., 2006). This process also estimates relatedness within the target sample, which is also important to consider in association analyses. Reference-projected PCs are estimated using the reference dataset, and then projected into the target sample, making them suitable for inclusion in prediction models (Chen et al., 2015).

#### User-Friendly Output

GenoPred generates detailed reports at both the sample- and individual-levels. These reports summarise input data processing metrics, ancestry inference results, and the polygenic scores. Individual-level reports provide ancestry inference and polygenic scores for a specific individual, presenting the polygenic scores on both relative and absolute scales to improve interpretability (Pain, Gillett, et al., 2022). The outputs of the pipeline are stored within a simple file structure that is easy to navigate. GenoPred also contains a series of R functions that use the pipeline configuration to read key pipeline outputs directly into R (R Core Team, 2015).

#### Configurable and Responsive

Users have the flexibility to trigger comprehensive analyses or request specific outputs of interest, avoiding unnecessary computational steps. For example, the user could request that the pipeline only performs quality control of the GWAS summary statistics, or that the pipeline only applies selected tuning parameters for a given polygenic scoring method. Modifications to input data or configurations prompt the pipeline to automatically update and rerun necessary steps, ensuring that outputs correspond to the latest data and research objectives. Various configuration parameters can be easily set by the user, allowing the user to adjust pipelines behaviour to suit their needs.

Technical documentation of each step in the pipeline is provided the Supplementary Data File 1 and online (https://opain.github.io/GenoPred/pipeline_technical.html).

### Application to UK Biobank

To demonstrate the simplicity, scalability and performance of the GenoPred pipeline, we applied it to calculate polygenic scores in the UK Biobank sample (Bycroft et al., 2018).

#### Configuration

We configured the GenoPred pipeline using three files (Figure 2). These directed GenoPred to compute polygenic scores within the UK Biobank using all 7 polygenic scoring methods across 13 GWAS summary statistics (Supplementary Table 1). The ‘config_file’ specifies the output directory, configuration file locations, and the polygenic scoring methods to be employed. The ‘target_list’ describes the target genetic dataset that polygenic scores should be calculated in. The ‘gwas_list’ describes the GWAS summary statistics that should be used to derive the polygenic scores.

In this study, using GenoPred release v2.2.5, we allowed 10 cores for polygenic scoring methodology and 50 cores for target samples, using the ‘cores_prep_pgs’ and ‘cores_target_pgs’ parameters respectively.

#### Execution

Once the configuration files have been prepared, a single line of code can be used to execute the full pipeline:

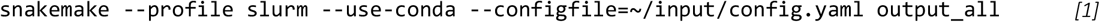

The ‘output_all’ command indicates which outputs of the pipeline are to be produced. In this case, it requests the final reports from the pipeline, which triggers all upstream steps of the pipeline (shown in Figure 1). A range of other commands can be used to produce intermediate or auxiliary outputs (Supplementary Data File 1). The ‘--configfile’ option tells the pipeline which configuration file to use. The ‘--profile slurm’ instructs the pipeline how to distribute the steps using the SLURM job scheduler for our HPC (King’s College London, 2022). Snakemake can interact with many different job schedulers or be run locally across cores using the ‘-j’ parameter. The ‘--use-conda’ option tells the pipeline to use Conda environments specified by each step in the pipeline.

**Figure 1.**
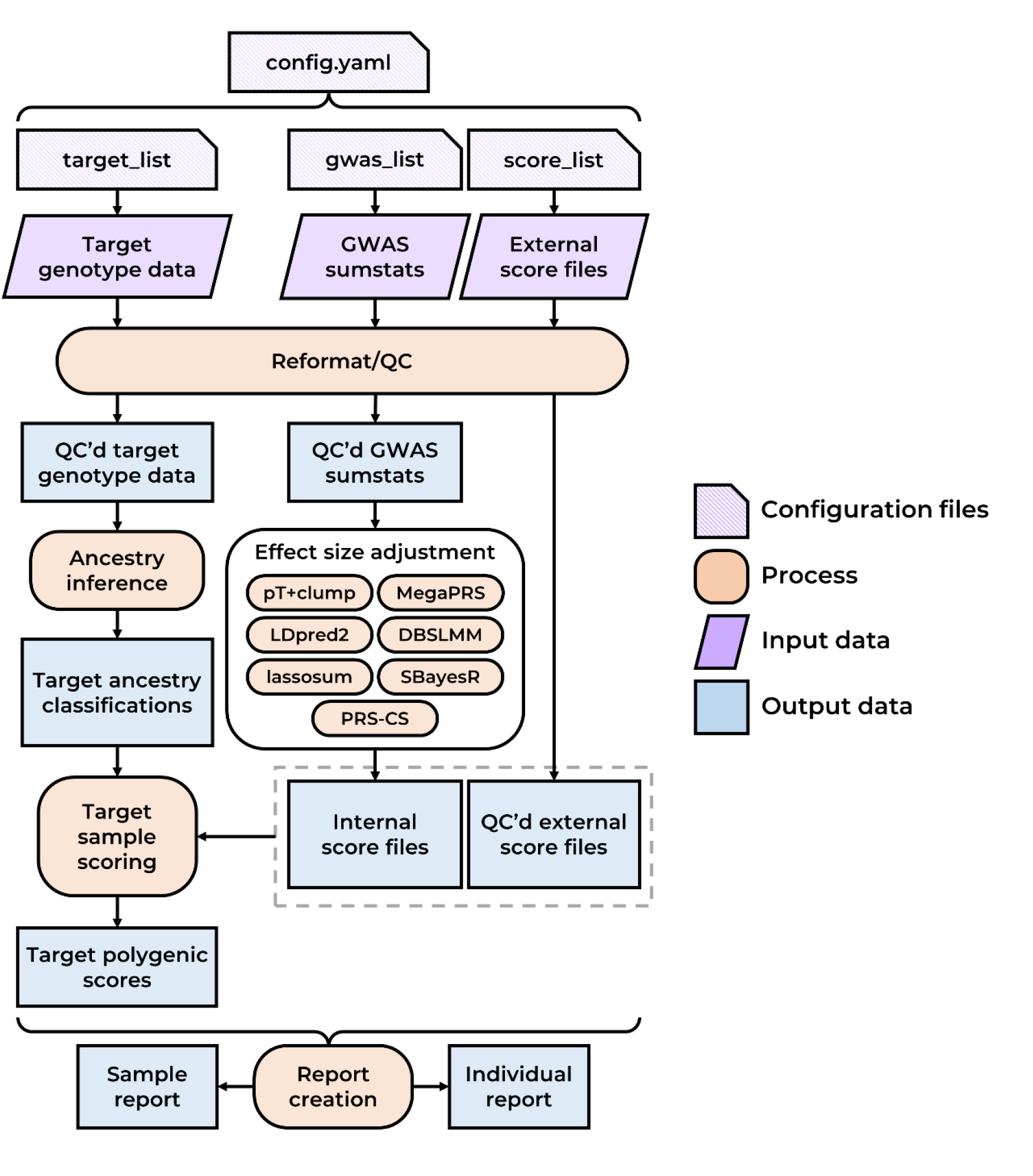
Simplified schematic diagram of the GenoPred pipeline. QC’d = Quality controlled.

**Figure 2.**
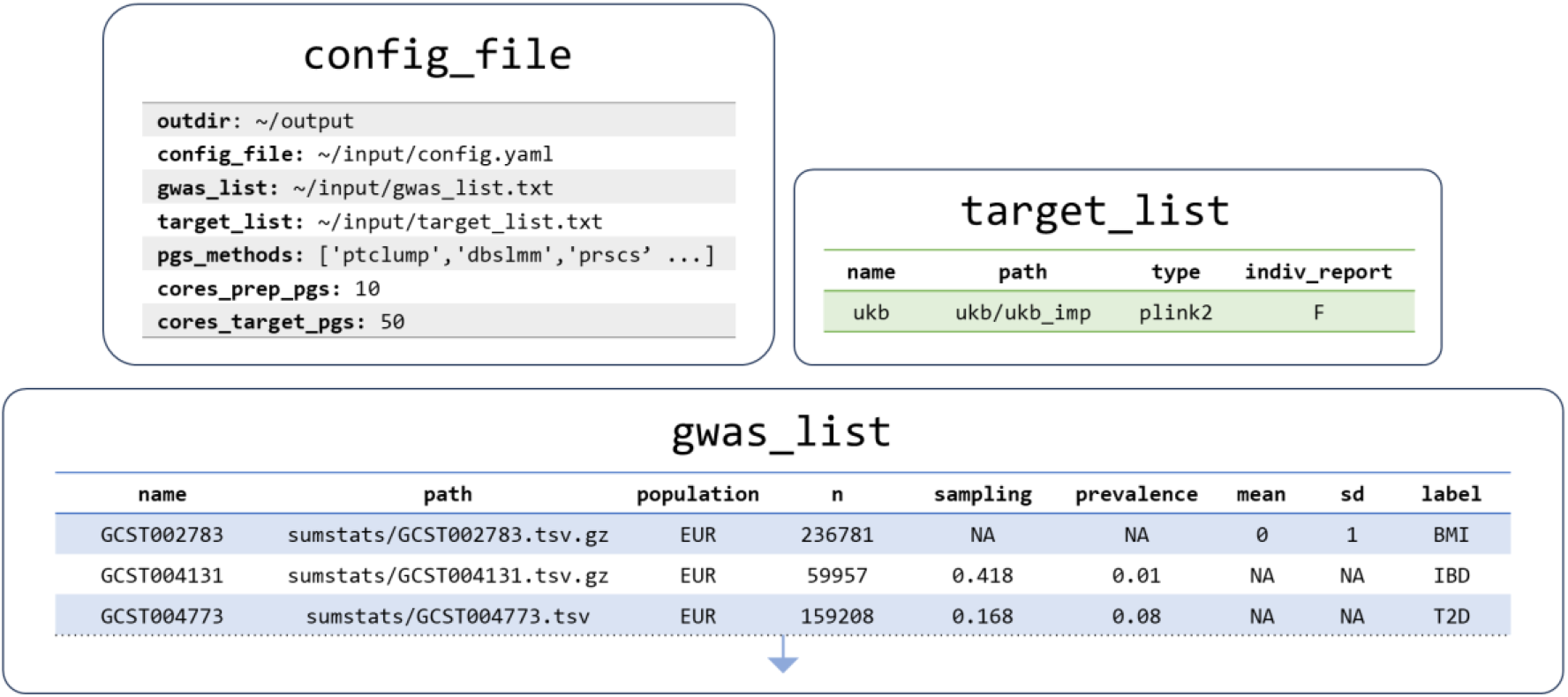
Configuration files instructing GenoPred pipeline to calculate polygenic scores in UK Biobank. Paths are simplified for clarity in this figure. Actual configuration files used are available in Supplementary Tables 2-4.

### Computational resources

The computational resources required vary at different stages of the pipeline and also depending on the pipeline configuration and input data. In this section we summarise the time required to run each step of the pipeline when using the UK Biobank configuration above. We used the full UK Biobank imputed genotype data in PLINK2 format as input (487,410 individuals, 9.94M variants).

When using the UK Biobank configuration described above the total computational wall time was 93.57 hours. When an HPC is available, and many steps can be executed in parallel, the time taken for the pipeline to finish is significantly reduced. Most of the time is spent applying polygenic scoring methodology to adjust GWAS effect sizes (85.5%), and then target scoring (7.5%) and target QC (6.3%) (Figure 3A). The time taken to perform target QC and target scoring is dependent on the number of individuals in the target datasets. The time taken for polygenic scoring methods to run is independent of the target datasets specified, but it will vary depending on which polygenic scoring methods are selected and the number of GWAS.

**Figure 3.**
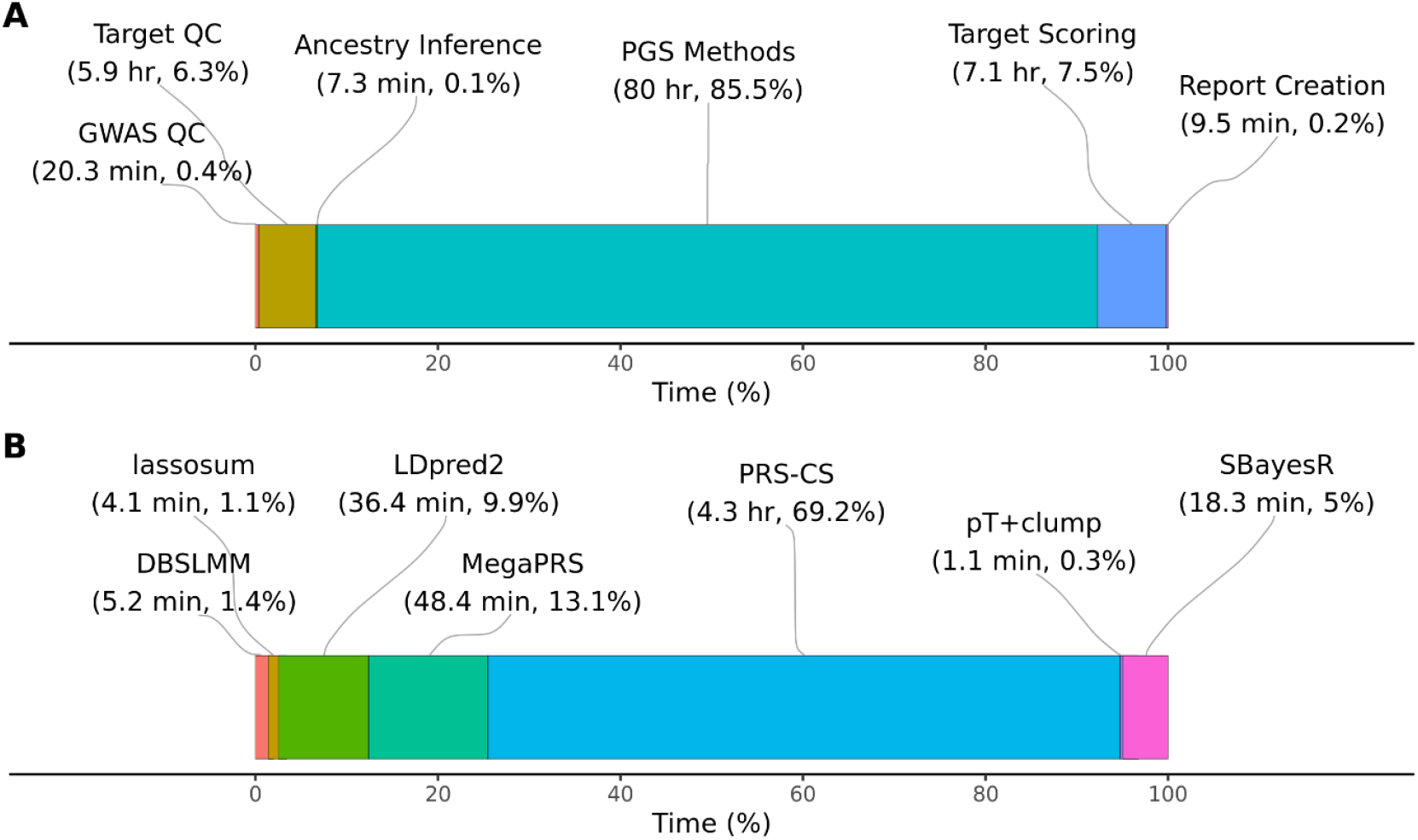
Wall-time for steps of the pipeline to complete using the UK Biobank configuration. A) Shows the total time for each step of pipeline, aggregating the polygenic scoring methods (labelled ‘PGS Methods’). B) Shows a breakdown of time for each polygenic scoring method, showing the average time per GWAS. QC = Quality control; PGS = polygenic scoring.

The time taken by each polygenic scoring method varies substantially (Figure 3B), and this may be an important factor when deciding the method/s to be used. Using the above configuration, GenoPred allocated 10 cores when running polygenic scoring methods. When using 10 cores, PRS-CS is the slowest of polygenic scoring methods, taking an average of 4.3 hours per GWAS. In contrast, all other methods can be run in less than 1 hour. The pT+clump, lassosum and DBSLMM methods are particularly efficient, taking less than 10 minutes.

### Performance

To demonstrate the performance of polygenic scores produced by the GenoPred pipeline, we estimated the association between polygenic scores and 14 relevant phenotypes, using univariate linear and logistic regression for continuous and binary outcomes respectively. Continuous outcomes and polygenic scores were standardised to have a mean of 0 and standard deviation of 1. The associations were estimated separately for each ancestral population in UK Biobank, as determined by the ancestry inference step of the pipeline (Supplementary Table 5). We used the same GWAS and UK Biobank phenotypes as a recent publication comparing polygenic scoring methodology for comparison (Monti et al., 2023), with some minor amendments (Supplementary Information 1). Sample sizes for each phenotype are in Supplementary Table 6.

Figure 4 shows the association results within the European subset of UK Biobank alone, to highlight relative performance of polygenic scoring methods. Figure 5 shows association results for continuous outcomes in all populations, to highlight relative association across populations. Supplementary Figure 1 shows results for binary outcomes in all populations. These plots show the association between polygenic scores and the relevant outcome. For example, for body mass index (BMI), a 1 standard deviation increase of the best performing polygenic score from LDpred2, is associated with a 0.3 standard deviation increase in observed BMI. Or for breast cancer, a 1 standard deviation increase of the best performing polygenic score from MegaPRS, is associated with an increase of 0.57 in the log(odds) of breast cancer (corresponding to an odds ratio of 1.77).

**Figure 4.**
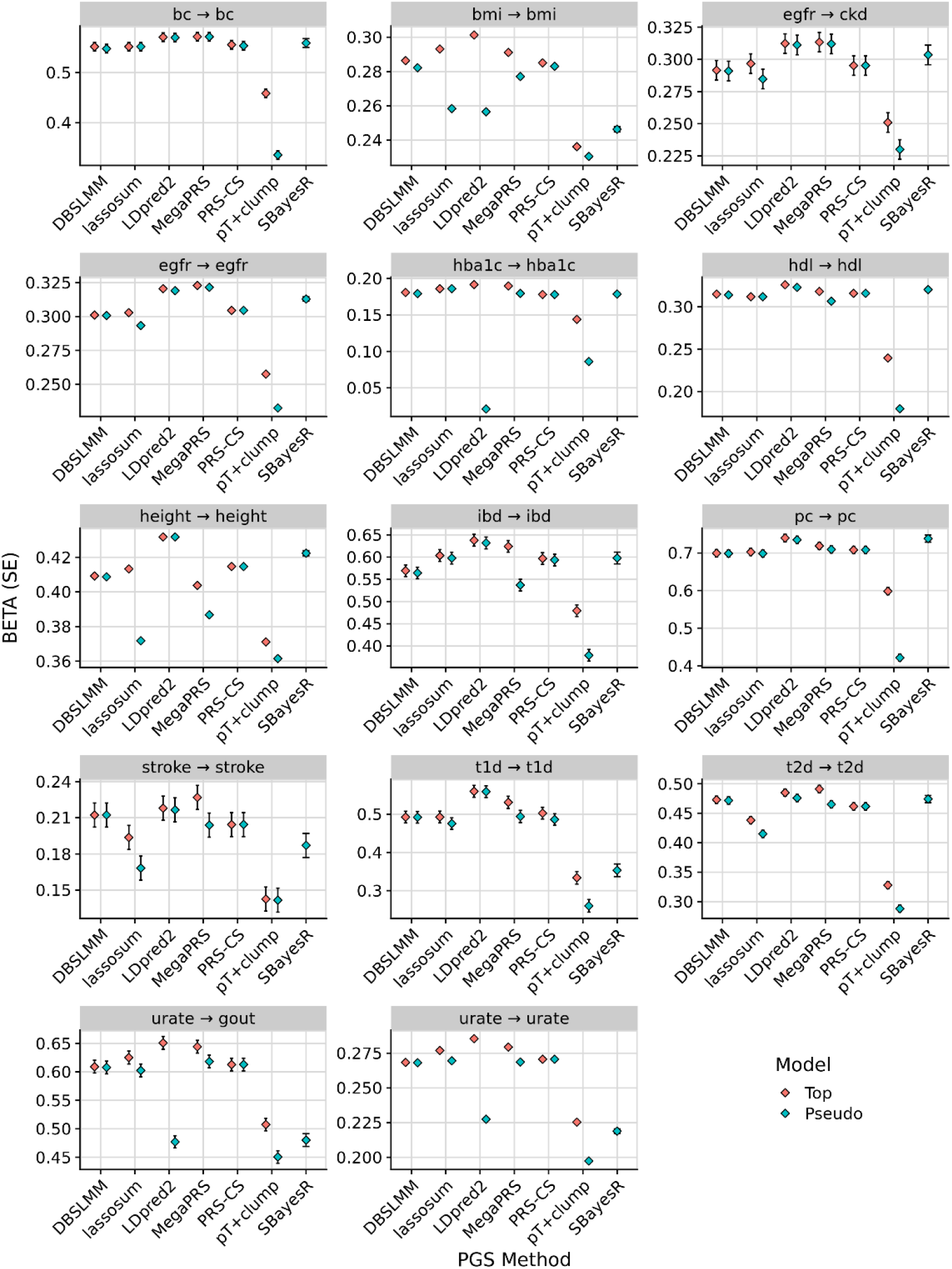
Association between polygenic scores and relevant phenotype data in the European subset of UK Biobank. For binary outcomes, the BETA corresponds to the log(odds ratio). The pseudo model refers to the polygenic score selected by each method’s pseudovalidation method, sometimes referred to as the ‘auto’ model. The pseudo model for the pT+clump method is a p-value threshold of 1. The top model is the polygenic score with the largest absolute correlation with the outcome. In the ‘egfr → ckd’ plot, the direction of associations was reversed to ensure the highest values correspond to the best performance in all plots. bc = breast cancer, bmi = body mass index, egfr = estimated glomerular filtration rate, ckd = chronic kidney disease, hba1c = hemoglobin A1c, hdl = high density lipoprotein, ibd = inflammatory bowel disease, pc = prostate cancer, t1d = type 1 diabetes, t2d = type 2 diabetes.

**Figure 5.**
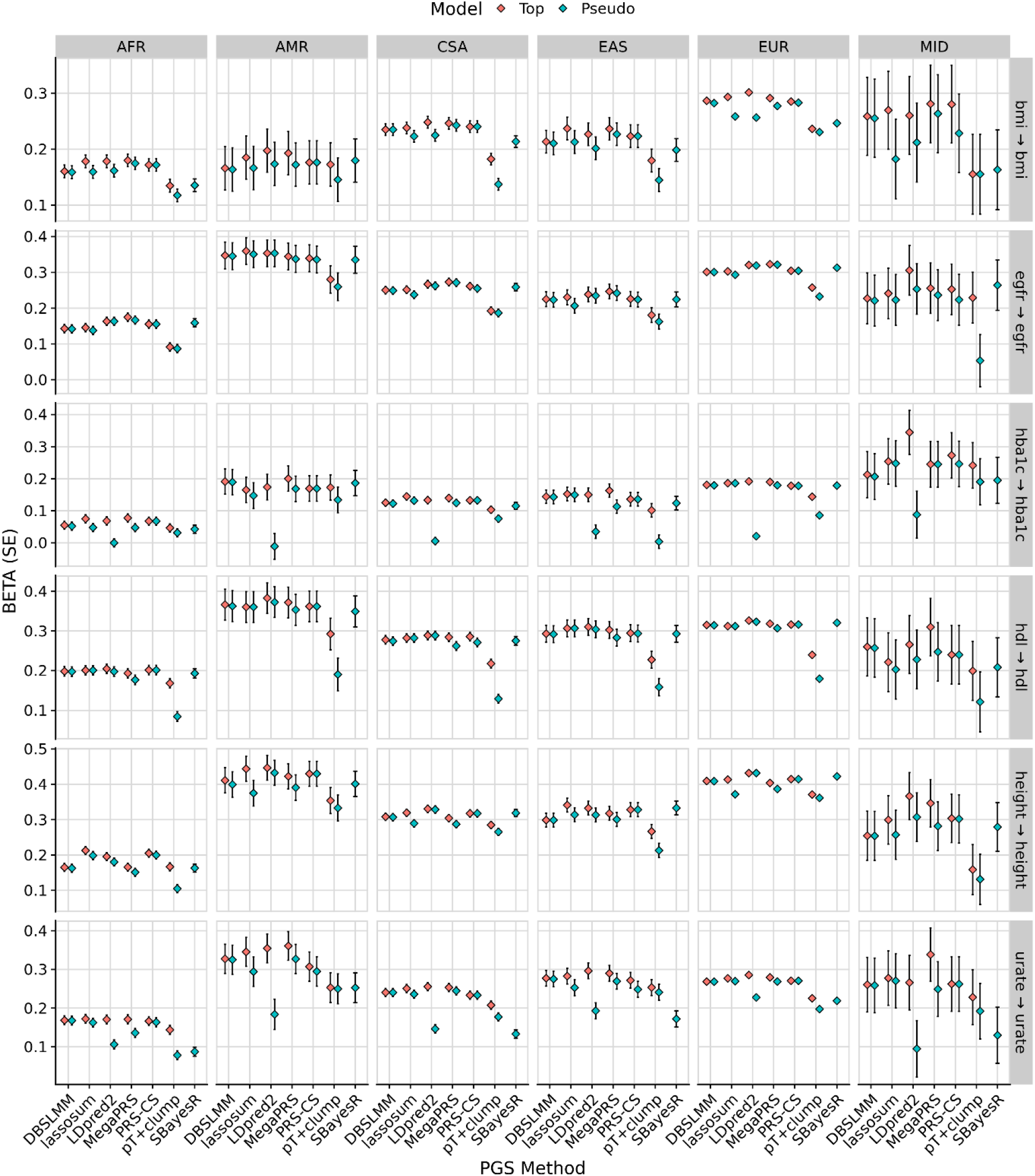
Association between polygenic scores and relevant continuous phenotype data in each UK Biobank population (AFR = African, AMR = Admixed American, CSA = Central and South Asian, EAS = East Asian, EUR = European, MID = Middle Eastern). The pseudo model refers to the polygenic score selected by each method’s pseudovalidation method, sometimes referred to as the ‘auto’ model. The pseudo model for the pT+clump method is a p-value threshold of 1. The top model is the polygenic score with the largest absolute correlation with the outcome. bmi = body mass index, egfr = estimated glomerular filtration rate, hba1c = hemoglobin A1c, hdl = high density lipoprotein.

Collectively, these results were congruent with the previous study by Monti and colleagues, with regard to the relative performance of each polygenic scoring method, and the absolute performance of the polygenic scores in UK Biobank.

## Discussion

The GenoPred pipeline represents a significant advance in the field of genetic epidemiology, particularly in the application and utility of polygenic scores. By integrating leading methodologies within a user-friendly and reproducible framework, GenoPred facilitates the generation of reliable and interpretable polygenic scores enabling a broader adoption of polygenic scoring in research and clinical settings where specialist computational and statistical expertise may not be available.

Our application of GenoPred to UK Biobank data underscores the pipeline’s efficiency, performance, and versatility. The results demonstrate GenoPred’s capability to handle large-scale datasets while maintaining high standards of quality control and ensuring the reproducibility of analyses. Importantly, by standardising the polygenic scoring process using reference genetic data, GenoPred enhances the interpretability of scores across diverse populations. This is crucial for the application of polygenic scores in global health, as it ensures that the benefits of genetic research are accessible to individuals of varied ancestries.

### Comparison to Existing Tools

To our knowledge, no other publicly available software offers such a comprehensive solution for polygenic scoring as the GenoPred pipeline. While many excellent tools exist for individual steps within the GenoPred pipeline, there is a notable absence of software that seamlessly integrates these steps. We will compare the GenoPred pipeline to several notable tools for polygenic scoring, highlighting the differences.

PRSice-2 is popular for its ease of use and computational efficiency (Choi & O’Reilly, 2019). A key feature of PRSice-2 is its ability to perform association analysis and output plots, a function currently not included in the GenoPred pipeline. However, PRSice-2 only includes the pT+clump polygenic scoring method, which underperforms compared to more recent methods (Pain et al., 2021). Furthermore, PRSice-2 is less flexible regarding GWAS or target sample data inputs, does not include target sample ancestry inference, and lacks reference-standardisation of polygenic scores, making it challenging to analyse and interpret scores in ancestrally diverse target samples. pgsc_calc is recently developed software primarily for target sample scoring using previously derived score files, stored within the Polygenic Score Catalog or elsewhere (Lambert et al., 2024). Like the GenoPred pipeline, pgsc_calc includes an ancestry inference module and standardises polygenic scores according to an ancestry-matched reference. Additionally, pgsc_calc offers a continuous correction to polygenic scores by regressing out reference-projected principal components, based on their relationship with polygenic scores in a reference dataset (Khera et al., 2019). This adjusts for population stratification within reference populations and returns polygenic scores for all individuals, not just those assigned to a reference population. However, pgsc_calc relies on previously derived scoring files, which may not exist for the GWAS of interest, or may not transfer well across samples due to missing genetic variants. In contrast, the GenoPred pipeline has the additional functionality to derive scoring files from GWAS summary statistics using the user’s choice of polygenic scoring method and genetic variants to be considered, substantially broadening its utility.

A range of other excellent tools for handling genetic data and polygenic scoring exist, such as the bigsnpr R package (Privé et al., 2018), PLINK2 (Chang et al., 2015), and LDAK (Zhang et al., 2021). However, these tools focus on specific tasks with a high degree of control and do not offer an end-to-end solution for polygenic scoring.

### Limitations

The current version of the GenoPred pipeline (v2.2.5) has two key limitations.

First, GenoPred assigns individuals to reference populations and standardises target sample scores according to the mean and standard deviation of the ancestry-matched population. While this solution is often sufficient for research contexts where downstream analyses are conducted in a population-specific manner, it leads to the exclusion of admixed individuals and those not well represented by the current reference populations. A potential solution to this limitation is the continuous ancestry adjustment of polygenic scores, as used in some previous studies in diverse populations (Khera et al., 2019), and implemented by the pgsc_calc software.

Second, GenoPred does not currently implement polygenic scoring methods that incorporate GWAS for a given outcome from multiple ancestral populations, such as PRS-CSx (Ruan et al., 2022) and BridgePRS (Hoggart et al., 2024). As genetic data becomes more widely available from diverse populations, methods incorporating GWAS from multiple populations are outperforming methods that only consider GWAS from a single population. Currently, to incorporate GWAS from multiple populations in GenoPred, users must either meta-analyse the GWAS and specify the majority population, or model polygenic scores from each population separately.

### Future Directions

As the field of genetic epidemiology continues to evolve, so too will the methodologies for polygenic scoring. GenoPred is well-positioned to adapt to these advances, thanks to its open-source nature and the active community of developers committed to its ongoing development. Future updates will aim to incorporate novel scoring methods, enhance support for multi-ancestry analyses, and improve the integration of functional genomic data to refine the predictive power of polygenic scores.

Moreover, the continued application of GenoPred across diverse research projects will generate valuable insights into its utility and limitations, guiding further refinements. Collaborations with clinical researchers will be particularly important for exploring the potential of polygenic scores to inform clinical decision-making and patient care.

### Conclusion

GenoPred fills a critical gap in the landscape of genetic research tools by providing a standardised, efficient, and accessible pipeline for polygenic scoring. Its development is a step forward in realizing the promise of personalized medicine and underscores the importance of making complex genetic analyses more accessible and interpretable. As we continue to enhance GenoPred and expand its capabilities, we look forward to its contribution to advancing our understanding of the genetic basis of diseases and traits and its application in improving human health.

## Data Availability

GWAS summary statistics used in this study were publicly available (see Supplementary Table 1). The UK Biobank data was accessed via project 82087 - For access, go to: https://www.UKBiobankiobank.ac.uk/enable-your-research/apply-for-access.

## Availability of supporting source code and requirements

Project Name: GenoPred pipeline V2

Project home page: https://opain.github.io/GenoPred/

Operating systems: Linux (64-bit) Programming language: R, python, bash

Other requirements (when recompiling): git, curl, wget, zip, gzip, build-essential, zlib1g-dev

License: GNU General Public License version 3.0 (GPLv3)

Any restrictions to use by non-academics: None

## Acknowledgments

We would like to thank Lucie Burgess, Michelle Kamp, Chris Lo, Julian Mutz, and Miryam Schattner for valuable feedback during the development of GenoPred pipeline. We would also like to thank Sam Lambert and Paul O’Reilly for their feedback on this manuscript.

OP is supported by a Sir Henry Wellcome Postdoctoral Fellowship [222811/Z/21/Z]. The funders had no role in study design, data collection and analysis, decision to publish, or preparation of the manuscript.

AAC is an NIHR Senior Investigator (NIHR202421). This is in part an EU Joint Programme - Neurodegenerative Disease Research (JPND) project. The project is supported through the following funding organisations under the aegis of JPND - www.jpnd.eu (United Kingdom, Medical Research Council (MR/L501529/1; MR/R024804/1)). This study represents independent research part funded by the National Institute for Health Research (NIHR) Biomedical Research Centre at South London and Maudsley NHS Foundation Trust and King’s College London.

## Conflict of Interest

OP provides consultancy services for UCB pharma. AAC reports consultancies or advisory boards for Amylyx, Apellis, Biogen, Brainstorm, Cytokinetics, GenieUs, GSK, Lilly, Mitsubishi Tanabe Pharma, Novartis, OrionPharma, Quralis, and Wave Pharmaceuticals. CML sits on the Myriad Neuroscience Scientific Advisory Board and is a Key Opinion Leader for UCB Pharma.

## Supplementary Information

### 1. Amendments to Monti et al. GWAS selection and phenotype definitions

We excluded the GWAS for Alzheimer’s disease as its sample overlaps with the UK Biobank target sample in this study. For Height, we used the Yengo et al. GWAS which excludes UK Biobank, allowing us to examine its performance in UK Biobank. The Height phenotype in UK Biobank was defined using the field 50 - Standing height. For RA, we used a more recent GWAS by Ishigaki et al (GCST90132223).

## Supplementary Figures

**Figure S1.**
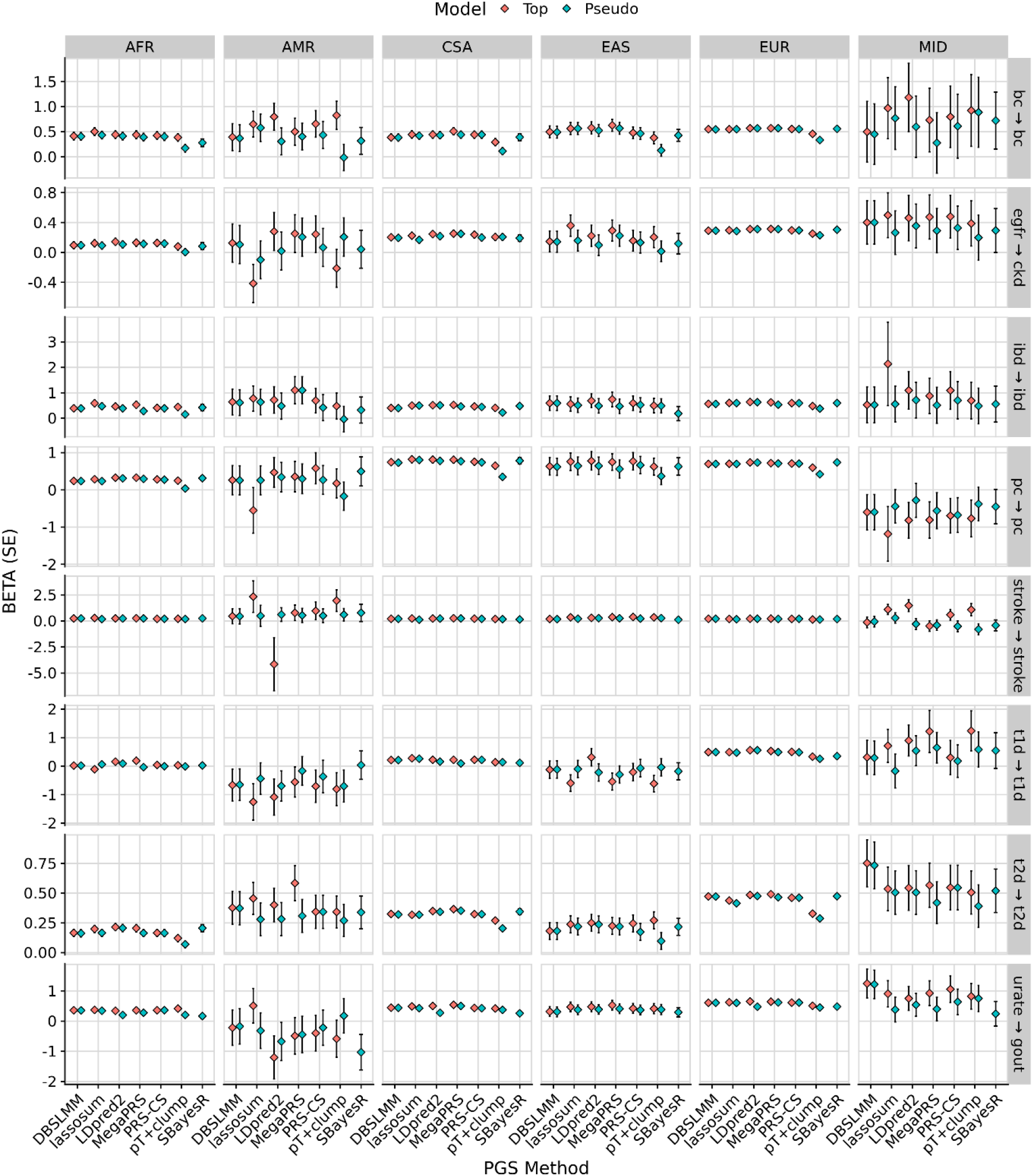
Association between polygenic scores and relevant binary phenotype data in each UK Biobank population (AFR = African, AMR = Admixed American, CSA = Central and South Asian, EAS = East Asian, EUR = European, MID = Middle Eastern). The pseudo model refers to the polygenic score selected by each method’s pseudovalidation method, sometimes referred to as the ‘auto’ model. The pseudo model for the pT+clump method is a p-value threshold of 1. The top model is the polygenic score with the largest absolute correlation with the outcome in each population. In the ‘egfr → ckd’ plot, the direction of associations was reversed to ensure the highest values correspond to the best performance in all plots. bc = breast cancer, ckd = chronic kidney disease, ibd = inflammatory bowel disease, pc = prostate cancer, t1d = type 1 diabetes, t2d = type 2 diabetes.

## Notes

### Author Declarations

UK Biobank data can be accessed via application, the data for this publication was accessed as part of projects 82087.

