## Supplementary Data 1 for "The GenoPred Pipeline: A Comprehensive and Scalable Pipeline for Polygenic Scoring"

### GenoPred Pipeline v2: Technical details

Written by Oliver Pain

### 1. Introduction

This document provides technical details of the GenoPred pipeline. The GenoPred pipeline automates the process of calculating polygenic scores. The pipeline aims to implement the current practises for polygenic scoring. See here more general information regarding the GenoPred pipeline.

### 2. Software

The pipeline, designed for both ease of use and reproducibility, is constructed using Snakemake. This workflow management system integrates a variety of scripts: R scripts, Python scripts, and bash scripts, providing a comprehensive toolkit. To ensure a seamless experience and maintain reproducibility, the entire pipeline operates within a Conda environment. We additionally provide docker and singularity containers to facilitate use in offline environments, and across operating systems. Moreover, for accessibility and collaboration, the pipeline has been made publicly available on GitHub, allowing users and developers alike to access, utilize, and contribute to its continuous improvement.

### 3. Workflow

The pipeline can be separated into two halves, reference data preparation and target data application. Reference data preparation refers to steps that only require reference data, including the reference genotype data, GWAS summary statistics, and externally generated score files. Target data application refers to steps that apply previously prepared reference data to the target datasets.

See here for more information on pipeline configuration.

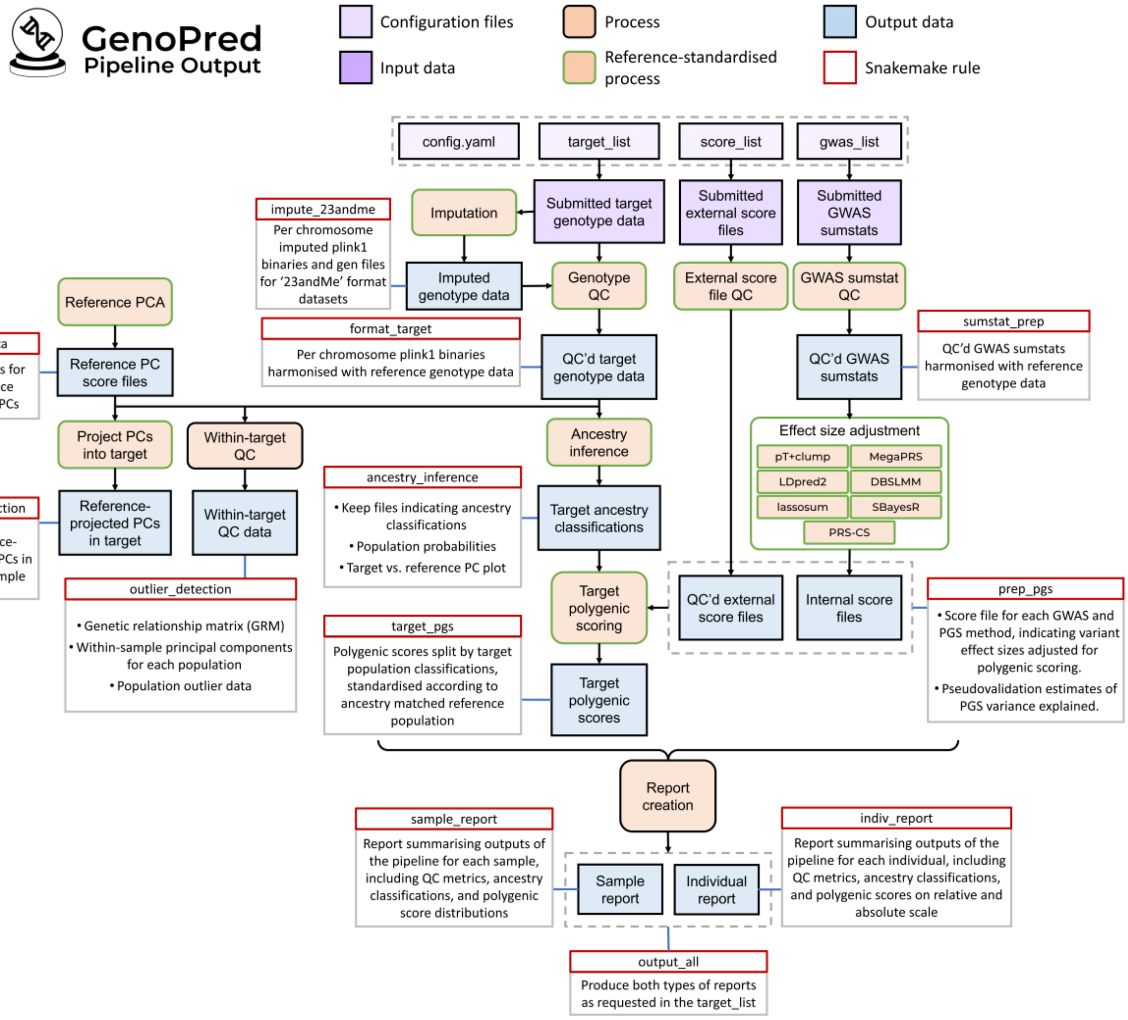

*Figure 1. GenoPred pipeline schematic. Shows input files, processes, outputs, and rules.*

### 4. Reference data preparation

#### 4.1. Reference genetic data

The default reference genotype data used by GenoPred is a previously prepared dataset, which is based on the 1000 Genomes Phase 3 (1KG) and Human Genome Diversity Project (HGDP) sample, restricted HapMap3 variants. This dataset contains 1204449 variants for 3313 individuals.

Users can provide their own reference data using the refdir parameter in the configfile. The reference data folder must have the following structure:

```
[refdir]
  ├── ref.chr[1-22].[pgen/pvar/psam] (plink2 genotype data with RSIDs in SNP column)
  ├── ref.chr[1-22].rds (SNP data - refer to default ref data for format)
  ├── ref.pop.txt (Population data for reference individuals - with header)
  ├── ref.keep.list (lists keep files for each population - columns pop and path - no
header)
  ├── keep_files
  │    └──[pop].keep (keep files for each population - no header)
  └── freq_files
       └──[pop]
           └──ref.[pop].chr[1-22].afrq (plink2 .afrq format)
```

**Note:** .psam, ref.pop.txt and keep_files must contain IID, and can optionally include FID information. The ID information must be consistent across these files.

#### 4.2. Reference PCA

Principal components analysis (PCA) using the reference genetic data is performed using the ref_pca.R script. First, PLINK v1.9 is used to retain variants with MAF > 0.05, missingness <0.02, and HWE p-value > 1e-6. Then, long range LD regions are removed (ref), and LD-pruning is applied using PLINK v1.9, using a 1000kb sliding window, with step size 5, and r2 threshold of 0.2 (–indep-pairwise 1000 5 0.2). Then PCA is performed using plink v2, storing SNP-weights for the first 6 PCs. The mean and SD of PCs within each reference population are calculated.

#### 4.3. GWAS sumstat QC

GWAS summary statistics, referred to as sumstats, submitted by the user are interpreted, formatted, and cleaned using the sumstat_cleaner.R script within the GenoUtils package. The GenoPred pipeline uses the default parameters for the script.

GWAS sumstats are read into R using the data.table::fread function, handling a range of column delimiters and compressions. The sumstat column names are interpreted and standardised using a dictionary. If sample size and allele frequency are reported for cases and controls, the total sample size and mean allele frequency are inserted.

The variants within the sumstats are then aligned to the 1KG+HGDP reference dataset. Strand ambiguous variants are removed. If chromosome number (CHR) and basepair position (BP) data is present in the sumstats, the genome build is identified through comparison of chromosome 22 data to a range of reference builds, considering the alleles of each variant (A1/A2) - IUPAC codes are used throughout, to allow for strand flips between the sumstats and reference. Then, sumstat variants are matched to reference variants using CHR, BP, and IUPAC data. If CHR and BP is not present, or genome build cannot be determined, RSIDs and IUPAC codes are used to match sumstat and reference variants. After matching with the reference, the reference allele frequency is inserted (REF.FREQ), using data from the 1KG+HGDP population matching that of the GWAS sample.

Variants in the summary statistics are then filtered according to the following criteria:

- Imputation quality (INFO): By default, variants with an INFO score < 0.9 are removed (if available in summary statistics).
- Minor allele frequency (MAF): MAF thresholds were set at 0.01. The script filtered variants below this threshold in either the GWAS sample (if available – FREQ) or reference data (REF.FREQ). Additionally, a MAF difference threshold (default 0.2) was used to exclude variants with substantial MAF discrepancies between GWAS and reference populations.
- P-Value: Variants with out-of-bound p-values (0 < P ≤ 1) were removed, ensuring statistical validity.
- Duplicate SNP ID: All variants with a duplicated SNP ID are removed.
- Sample Size: Variants with sample size >3SD from the median are removed (if per variant sample size is available in summary statistics).

Finally, the script inserts a BETA coefficient column it is not present already, calculating as log(OR), or using signed Z-scores, sample size and allele frequency estimates (ref). The standard error (SE) is also inserted if it is not present, subsequently removing variants with a SE equal to zero. If the SE is available, there is also a check whether P values have been adjusted using genomic control – If genomic control is detected, the p-value is recomputed using Z scores (BETA/SE).

#### 4.4. Adjustment of GWAS effect sizes

The GenoPred pipeline integrates various polygenic scoring methods for adjusting effect sizes in GWAS summary statistics. It uses a reference-standardized approach, adjusting GWAS effect sizes based on an ancestry-matched reference sample, without considering specific target sample characteristics.

##### 4.4.1. Single Ancestry Methods

The GenoPred pipeline currently only implements PGS methods intended for GWAS based on a single ancestral population. When using GWAS based on mixed ancestry individuals, we suggest specifying the population that matches most individuals in the GWAS.

Table 1 provides a summary of each approach and its implementation in GenoPred.

###### 4.4.1.1. DBLSMM

DBSLMM, a Bayesian shrinkage method, is implemented using the script pgs_methods/dbslmm.R. We use the 'tuning' version, SNP-based heritability parameter is varied by factor of 0.8, 1, and 1.2. When using a factor 1, this corresponds to the 'default' version of DBSLMM. The SNP-based heritability (on the liability scale for binary outcomes) is estimated using LD-score regression, using LD scores released by pan-UK (link). GenoPred clones DBSLMM (d158a144dd2f2dc883ad93d0ea71e8fc48e80bd3) from GitHub, downloads the precompiled software binary provided by DBSLMM authors, and uses the 1KG+HGDP reference for LD estimation based on the super population of the GWAS sample. LD block data is only provided for EUR, EAS and AFR populations - If the GWAS are from CSA, AMR, MID populations, EUR LD block data is used.

###### 4.4.1.2. lassosum

lassosum, an R package, applies the LASSO method for constructing PGS, is implemented using the script pgs_methods/lassosum.R. It uses a range of lambda hyperparameters and offers a pseudovalidation feature to deduce the optimal lambda value, eliminating the need for external testing data. GenoPred uses lassosum v0.4.5, with the default range of lambda values, and the 1KG+HGDP reference for LD estimation, matching the population of the GWAS sample. LD block data for EUR, EAS and AFR are provided by lassosum - If the GWAS are from CSA, AMR, MID populations, EUR LD block data is used.

###### 4.4.1.3. LDpred2

LDpred2, part of the bigsnpr R package, operates in ‘inf’, ‘grid’, and ‘auto’ modes. The ‘inf’ mode is better suited for highly polygenic traits, whereas ‘grid’ and ‘auto’ adjust effect sizes using various hyperparameters, including SNP heritability. In GenoPred, LDpred2 is run using the script pgs_methods/ldpred2.R. GenoPred uses bigsnpr v1.12.2, with the default LDpred2 grid search, and recommended GWAS quality control check. GenoPred employs LDpred2’s precomputed LD matrices based on the European individuals from the UK Biobank, and it is applied only to GWAS based on a EUR population. If the SNP-h2 estimated using LDSC is <0.05, the SNP-heritability used by LDpred2 is set to 0.05.

###### 4.4.1.4. MegaPRS

MegaPRS uses a range of priors (lasso, ridge, bolt, BayesR) for SNP effects, run via LDAK software. In GenoPred, MegaPRS is run using the script pgs_methods/megaprs.R. The GenoPred pipeline uses LDAK v5.1, with the BLD-LDAK heritability model, where SNP variance is dependent on allele frequency, LD, and functional annotations. It includes a pseudovalidation approach. GenoPred employs the 1KG+HGDP reference for LD estimation, matching the population of the GWAS sample.

###### 4.4.1.5. PRS-CS

PRS-CS, a Bayesian method using a continuous shrinkage prior, specifies a range of global shrinkage parameters (phi), generating multiple sets of genetic effects for polygenic scoring. Its ‘auto’ model estimates the optimal parameter directly from GWAS summary statistics, negating the need for an external dataset. In GenoPred, PRS-CS is run using the script pgs_methods/prscs.R. GenoPred specifies four phi parameters (1e-6, 1e-4, 1e-2, 1) and the auto model. By default, GenoPred uses the PRS-CS provided 1KG-derived LD matrix data, matching the population of the GWAS sample. The user can select the UKB-derived LD matrix data to be used using the prscs_ldref parameter in the configfile. 1kg is used by default as PGS based on Yengo et al. sumstats performed significantly better in the OpenSNP target sample, when using the 1KG reference data (this may differ for other GWAS).

###### 4.4.1.6. pT+clump

In GenoPred, pT+clump is run using the script pgs_methods/ptclump.R. This method conducts LD-based clumping (using PLINK v1.9, r2 = 0.1; window = 250kb) and shrinks effects to zero across a range of p-value thresholds (default: 1e-8, 1e-6, 1e-4, 1e-2, 0.1, 0.2, 0.3, 0.4, 0.5, 1). To address long-range LD in the MHC/HLA region (Chr6:28-34Mb), only the most associated variant is used for polygenic scoring. GenoPred uses the 1KG+HGDP reference for LD estimation, matching the population of the GWAS sample.

###### 4.4.1.7. SBayesR

In GenoPred, SBayesR is run using the script pgs_methods/sbayesr.R. Implemented with GCTB software, SBayesR is a Bayesian method that uses GWAS summary statistics to estimate key parameters. GenoPred uses GCTB v2.03 with LD matrices from the GCTB authors, calculated using European individuals in the UK Biobank, restricting its application to GWAS based on EUR populations. GenoPred uses the robust parameterisation option in SBayesR.

| Method | Software | PubMed ID | Pseudovalidation Option | Parameters | MHC Region | LD Reference | CPU usage* | Memory Usage* |
|---|---|---|---|---|---|---|---|---|
| DBSLMM | DBSLMM | 32330416 | Yes (only option) | SNP-heritability estimated using LD Score Regression (on liability scale for binary outcomes) | Not excluded | Population-matched 1KG+HGDP | 40 seconds | 450 Mb |
| lassosum | lassosum R package | 28480976 | Yes | s = 0.2, 0.5, 0.9, 1; lambda [A] = exp(seq(log(0.001), log(0.1), length.out=20)) | Not excluded | Population-matched 1KG+HGDP | 10 seconds | 400 Mb |
| LDpred2 | bigsnpr R package | 33326037 | Yes | Grid includes 126 combinations of heritability and non-zero effect fractions (p). | Not excluded | EUR UKB (LDpred2-provided) | 3 minutes | 500 Mb |
| MegaPRS | LDAK | 34234142 | Yes | Fits lasso, ridge, bolt and BayesR models, with a total of 148 sets of hyperparameters | Not excluded | Population-matched 1KG+HGDP | 5 minutes | 450Mb |
| PRS-CS | PRS-CS | 30992449 | Yes | phi = 1e-6, 1e-4, 1e-2, 1, auto | Not excluded | Population-matched UKB (PRS-CS provided) | 35 minutes | 350 Mb |
| pT+clump | PLINK | 25722852 | No | p-value thresholds: 1e-8, 1e-6, 1e-4, 1e-2, 0.1, 0.2, 0.3, 0.4, 0.5, 1; Clumping: r2 = 0.1; window = 250kb | Only top variant retained | Population-matched 1KG+HGDP | 5 seconds | 100 Mb |
| SBayesR | GCTB | 31704910 | Yes (only option) | NA | Excluded (as recommended) | EUR UKB (GCTB-provided) | 3 minutes | 500 Mb |

[A] lassosum lambda values described using R code.

[B] See here for full benchmark information.

##### 4.5. External score file QC

This step harmonises externally derived score files with the reference genetic data. It is performed using the external_score_processor.R script. The script requires the PGS Catalogue header format. First the script checks whether the PGS file contains comments in the header, and if it does, records the reported genome build. It then reads in the score files, updating retaining the PGS catalogue harmonised columns if they are present. The variants within the score file are then aligned to a reference dataset - This is the same process used to harmonise GWAS summary statistics (Section 4.3), except the script will use the reported genome build if present.

#### 5. Target data application

The user can provide individual-level target genotype data in several formats, including PLINK1, PLINK2, VCF, BGEN, or 23andMe. PLINK1, PLINK2, VCF and BGEN datasets are assumed to have already undergone genotype imputation and can include 1 or more individuals. 23andMe format data contains data for a single individual and is assumed to be unimputed. Therefore, prior to reference harmonisation, 23andMe data undergoes imputation and is converted to PLINK1 format within the GenoPred pipeline.

##### 5.1. Target genotype imputation

Genotype imputation is only performed for target datasets in the format of 23andMe. This functionality is included for polygenic scores to be calculated for 23andMe datasets. GenoPred is not intended for large-scale genotype imputation – Instead we would recommend using free and publicly available imputation servers, which leverage large-scale haplotype reference panels. GenoPred requires target datasets in formats other than 23andMe to have been imputed in advance, or to data that does not require imputation (such as whole genome sequence data).

Genotype imputation in GenoPred is performed using the 23andMe_imputer.R script, which is largely based on the code written by Lasse Folkersen for the website Impute.Me. In GenoPred, genotype imputation is performed using shapeit and impute2, using the 1KG Phase 3 reference. The imputed data is then converted to PLINK1 binary format, using PLINK2 and a –hard-call-threshold of 0.1.

##### 5.2. Target genotype QC

Target genotype QC is performed using the format_target.R script. Each chromosome of the input data is processed in parallel. Initially, reference and target sample variant data are read into R. Then the genome build of the target data is identified through comparison of CHR, BP, and IUPAC information in the reference data. Then, variants in the target data are matched to those in the reference data, identifying variants that are strand flipped, have mismatch RSIDs and duplicate variants. PLINK2 is then used create PLINK2 format target genotype data, extracting variants in common with the reference, updating IDs, and managing any needed strand flips. If input target genotype data is in VCF or BGEN format, either the PLINK –vcf-min-gq 10 or –hard-call-threshold 0.1 parameters are applied respectively. Finally, reference variants that are not present in the target data are inserted as missing, to allow reference allele frequency-based imputation during downstream polygenic scoring.

#### 5.3. Ancestry Inference

Target samples then undergo ancestry inference, using the Ancestry_identifier.R script, estimating the probability that each target individual matches each reference population (AFR = African, AMR = Admixed American, EAS = East Asian, EUR = European, CSA = Central and South Asian, MID = Middle Eastern). Population membership was predicted using a reference trained elastic net model consisting of the first six reference-projected genetic principal components. Principal components were defined in the reference dataset using variants present in the target dataset with a minor allele frequency >0.05, missingness <0.02 and Hardy-Weinberg p-value >1×10-6 (if target sample size <100, then only missingness threshold is applied in the target). LD pruning for independent variants is then performed in PLINK after removal of long-range LD regions (ref), using a window size of 1000, step size of 5, and r2 threshold of 0.2. The A multinomial elastic net model predicting super population membership in the reference is derived in using the glmnet R package, with model performance assessed using 5-fold cross validation. The reference-derived principal components are then projected into the target dataset, and the reference-derived elastic net model is used to predict population membership in target. By default, target individuals are assigned to a population if the predicted probability was >0.95, but the user can modify this threshold using the ancestry_prob_thresh parameter in the config file.

#### 5.4. Within-target QC

GenoPred can also perform several quality control functions within the target sample, using script outlier_detection.R. Note, this step is not reference-standardised, making it less suitable for prediction modelling, but is still useful for accounting for population stratification when inference is the aim.

If the user doesn't provide a list of unrelated indiviuals using the unrel column in the target_list, the relatedness across the full target sample is estimated using the KING estimator in PLINKv1.9. A list of unrelated individuals is defined using a threshold of >0.044. Then within each population with at with at least 100 individuals (assigned by the ancestry inference step), PCA is performed to produce within-sample PCs (as opposed to reference-projected PCs) - These may capture target sample specific structure that reference-projected PCs will not, such as batch effects. PCA was applied after LD-pruning and removal of long-range LD regions (ref). Using the top 10 within-sample PCs, outliers are detected using the distance to k-means centroids. PCs and outlier thresholds are defined using a maximum of 1000 unrelated individuals, and then projected onto the full target sample. The number of clusters within each population was defined using the NbClust R package. Individuals were removed if their distance from any one centroid was greater than Q3+30×IQR, where Q3 is the third quartile, and IQR is the interquartile range. This threshold is arbitrary, but was decided after visual inspection of centroid distance distributions in UK Biobank and seems to generalise fairly well.

#### 5.5. Target scoring

This step calculates scores in the target sample, based on scoring files from the PGS methods, and from the reference PCA. It is carried out using the target_scoring.R script. It is performed separately for each target sample population. Scoring is performed using PLINK2 using the –read-freq function to impute missing genotypes using the ancestry-matched reference allele frequency. The scores are computed for each chromosome, then multiplied by the number of non-missing variants to get the sum score, and then added across chromosomes. Finally, the target sample scores are standardised according to the mean and SD of the score in an ancestry-matched reference.

#### 5.6. Report creation

##### 5.6.1. Individual-level

This step creates an .html report summarising the pipeline outputs for each individual in the target sample. It simply reads in pipeline outputs, and then tabulates and plots them. The only analysis it performs is the conversion of polygenic scores onto the absolute scale. It uses a previously published method. The estimate of the PGS R2 come from the lassosum pseudovalidation analysis, and the distribution in the general population is provided by the user in the prev, mean and sd columns of the gwas_list. Note. It does not convert PGS from externally derived polygenic scores onto the absolutes scale.

##### 5.6.2. Sample-level

This step creates an .html report summarising the pipeline outputs for each target sample. It simply reads in pipeline outputs, and then tabulates and plots them.
